# TMS–EEG-derived excitation/inhibition ratio as a diagnostic biomarker for major depressive disorder

**DOI:** 10.64898/2026.01.22.26344599

**Authors:** Elena Ukharova, Isabella O’Meeghan, Ida Granö, Sabin Sathyan, Emi Iizuka, Casper Tillander, Noora Kainulainen, Paula Partanen, Maria Vesterinen, Satu Palva, J. Matias Palva, Timo Roine, Juha Gogulski, Zafiris J. Daskalakis, Daniel M. Blumberger, Risto J. Ilmoniemi, Pantelis Lioumis

## Abstract

Major depressive disorder (MDD) is characterized by both physiological and psychological changes that impact daily function and quality of life. Despite advancements in treatment approaches, clinical outcomes remain variable, highlighting the need for reliable biomarkers to guide treatment selection and to monitor treatment responses. Transcranial magnetic stimulation combined with electroencephalography (TMS–EEG) allows for the characterization of local cortical excitation and inhibition through TMS-evoked potentials (TEPs). However, interindividual variability and various artifacts have previously limited the utility of TEPs for biomarker development.

We applied single-pulse TMS to the left dorsolateral prefrontal cortex (L-DLPFC), and recorded EEG responses in 28 individuals with MDD and 8 healthy controls. An individualized mapping procedure identified stimulation sites and intensities that minimized artifacts and ensured early TEP (<60 ms) deflections exceeded the predefined threshold of 6 μV. We quantified the ratio between the second and first peak-to-peak amplitudes of the TEP, as excitatory and inhibitory mechanisms have been identified to contribute to these peaks.

The TEP peak ratio differed significantly between MDD participants and healthy controls (*p* = 0.003), and correlated with depression symptom severity quantified with the Quick Inventory of Depressive Symptomatology Self-Report (*r* = 0.459, *p* < 0.05), Ruminative Response Scale (*r* = 0.656, *p* < 0.001), Patient Health Questionnaire (*r* = 0.368, *p* < 0.05), and Positive Valence Systems Scale (*r* = 0.332, *p* < 0.05). These correlations were not observed when an anatomy-based targeting method (Beam F3) was applied with the conventional stimulation intensity of 120% of the resting motor threshold.

Our findings highlight the utility of a novel individualised mapping technique in capturing high-quality early TEPs through a real-time EEG readout for minimisation of artifacts and ensuring a minimum TMS-evoked response. The altered TEP peak ratio adds to the growing body of evidence that the excitation/inhibition imbalance plays a role in MDD neurophysiology and serves as a promising candidate biomarker that warrants further investigation.

## Introduction

Currently, depression is diagnosed largely on the basis of subjective symptom questionnaires and clinical interviews. With more than 20 pharmacological and non-pharmacological interventions approved for MDD, many patients undergo a long course of trial and error process, possibly leading to significant delays in achieving treatment response and remission (Leuchter et al., 2009). Fewer than 50% of individuals with depression respond to the first antidepressant, and only 30% achieve full remission (Trivedi et al., 2006). Identifying objective biomarkers that can predict responses and guide personalised treatment approaches could significantly improve clinical outcomes.

Increasingly, psychiatric conditions are conceptualized as disorders of brain networks (Gorman, 1996). In MDD, the dorsolateral prefrontal cortex (DLPFC) and the subgenual anterior cingulate cortex (sgACC) are considered key nodes, given their observed abnormalities in depression and their modulation in various depression-treatment modalities (Koenigs and Grafman, 2009). Metabolic hyperactivity in the sgACC and its anticorrelated functional connectivity with the left DLPFC (L-DLPFC) have been found to be predictive of the response to the repetitive transcranial magnetic stimulation (rTMS) treatment, supporting the relevance of functional sgACC–DLPFC connectivity for the selection of the rTMS stimulation targets in depression (Cash et al., 2021; Fox et al., 2012; Weigand et al., 2018).

Concurrent TMS and EEG (TMS–EEG) enable direct evaluation of cortical properties, showing promise as a tool for identification of biomarkers of psychiatric conditions (Cao et al., 2021; Kallioniemi and Daskalakis, 2022). Changes in TMS-evoked potentials (TEPs) have been linked to depression diagnosis and symptom severity. In particular, when stimulating the L-DLPFC, MDD patients show larger amplitudes of peaks occurring at 45, 60, and 100 ms post-stimulus, referred to as N45, P60, and N100, have been shown to be larger than in healthy controls (Voineskos et al., 2019). Additionally, P60 and N100 amplitudes have been correlated with depression symptom severity (Cash et al., 2021; Fox et al., 2012; Li et al., 2021; Weigand et al., 2018). Increased cortical excitation has also been reported, evidenced by increased global mean field amplitude area under the curve (GMFA–AUC; Voineskos et al., 2019). Baseline P30, N45, P60, and N100 amplitudes have also correlated with or been predictive of the degree of response to theta-burst stimulation (TBS; Dhami et al., 2023), rTMS (Eshel et al., 2020) and magnetic seizure therapy (MST; Sun et al., 2016).

While these preliminary findings show promise towards identifying robust TMS–EEG biomarkers in MDD, several methodological limitations constrain the interpretation and reproducibility of TMS-evoked potentials. Early TEP components that are considered to reflect local cortical reactivity (Ilmoniemi et al., 1997) are markedly impacted by artifacts, and demand individualized targeting and rigorous artifact control for reliable recording. Conversely, the later components are more unspecific with regard to stimulation, and may include auditory and somatosensory confounds (Conde et al., 2019; Hernandez-Pavon et al., 2023; Tremblay et al., 2019). Additionally, the early TEP components are connected to the local NMDA- and GABA-A-ergic activity, which potentially allow for the development of biomarkers that isolate the local excitation/inhibition balance (Darmani and Ziemann, 2019).

For identifying the L-DLPFC stimulation site, previous work on TMS–EEG biomarkers in depression has relied on Talairach coordinates (Voineskos et al., 2019), MNI coordinates (Dhami et al., 2023, 2020), and EEG electrode positions (Biermann et al., 2022; Sun et al., 2016). These methods, despite standardised methodology, neglect the effect of interindividual differences in head anatomy, scalp-to-cortex distance, cortical morphology, and gyrification. Scalp-based methods can miss the DLPFC by 2 cm when compared to using an individual’s MRI (Ahdab et al., 2010). Furthermore, even if identical anatomical areas are stimulated, this does not necessarily guarantee identical functional correspondence due to interindividual variability in structure-function relationships (Casali et al., 2010). Additionally, previous work has utilized fixed coil orientations (Biermann et al., 2022; Dhami et al., 2023, 2020; Sun et al., 2016; Voineskos et al., 2019), disregarding individual differences in gyral morphology of the DLPFC.

Stimulation intensity is typically defined relative to the resting motor threshold (RMT), without accounting for individual anatomical differences or DLPFC reactivity (Biermann et al., 2022; Dhami et al., 2023, 2020; Sun et al., 2016; Voineskos et al., 2019). However, electric-field (E-field) modeling and monitoring reveal large inter-individual differences in cortical dosing at a given RMT defined by stimulator output. Consequently, the conventional 120% RMT approach risks substantial over- or under-dosing, underscoring the need for individualized E-field monitoring (Caulfield et al., 2021).

While most prior TMS–EEG studies in depression have focused on absolute peak amplitudes (Ziemann et al., 2026), isolated deflections provide only a limited description of cortical reactivity. In contrast, relationships between consecutive components reflect the temporal integration of excitation and inhibition and may therefore be more informative about complex cortical dynamics (Chen et al., 2025; Rissanen et al., 2026). Here, we introduce a novel approach for quantifying early local cortical activity based on the ratio of the first two prominent peak-to-peak amplitudes within the 10–60 ms window. This approach also normalizes the resulting measures across individualized stimulation targets and intensities (Ukharova et al., 2025). As specific TEP components have been linked to both excitatory and inhibitory neurotransmitter systems, we interpret these ratios as an index of local excitation–inhibition balance.

The present study focused on alterations in local cortical excitability (Casarotto et al., 2022; Lioumis and Rosanova, 2022; Tervo et al., 2022) via early TEP responses (10–60 ms). We compared TEPs obtained from MDD patients and healthy controls through standardised methodology (Beam F3; Beam et al., 2009), and contrasted these with the results obtained with individualised mapping of TMS–EEG guided by brain anatomy, functional connectivity, and real-time responses, as demonstrated in our recent study (Ukharova et al., 2025). To investigate whether the newly introduced peak ratio and previously established N100 amplitude biomarker scale with depressive symptom severity, we correlated TEP-derived measures with behavioral scores from the Patient Health Questionnaire (PHQ), Quick Inventory of Depressive Symptomatology Self-Report (QIDS-SR), Ruminative Response Scale (RRS), and Positive Valence Systems Scale (PVSS), testing the hypothesis that alterations in local excitation–inhibition balance and inhibitory processing within the L-DLPFC are directly related to the clinical expression of depression. Finally, we also examined whether TMS–EEG measures differ systematically between MDD patients and healthy controls.

## Methods

### Participants

28 MDD patients and 8 healthy controls (demographics, comorbidities, and medication statuses are presented in Table 1) were recruited from the participant pool of the larger PlaStim (Plasticity Stimulation in the Treatment of Anhedonia) study of Wellcome Leap Multichannel Psych Program (CA, USA). The sample size discrepancy reflected prioritization of clinical data collection within project timelines. The study was approved by the Helsinki University Hospital Ethics Committee (HUS/3043/2021). Written informed consent was obtained from all participants.

**Table 1.**
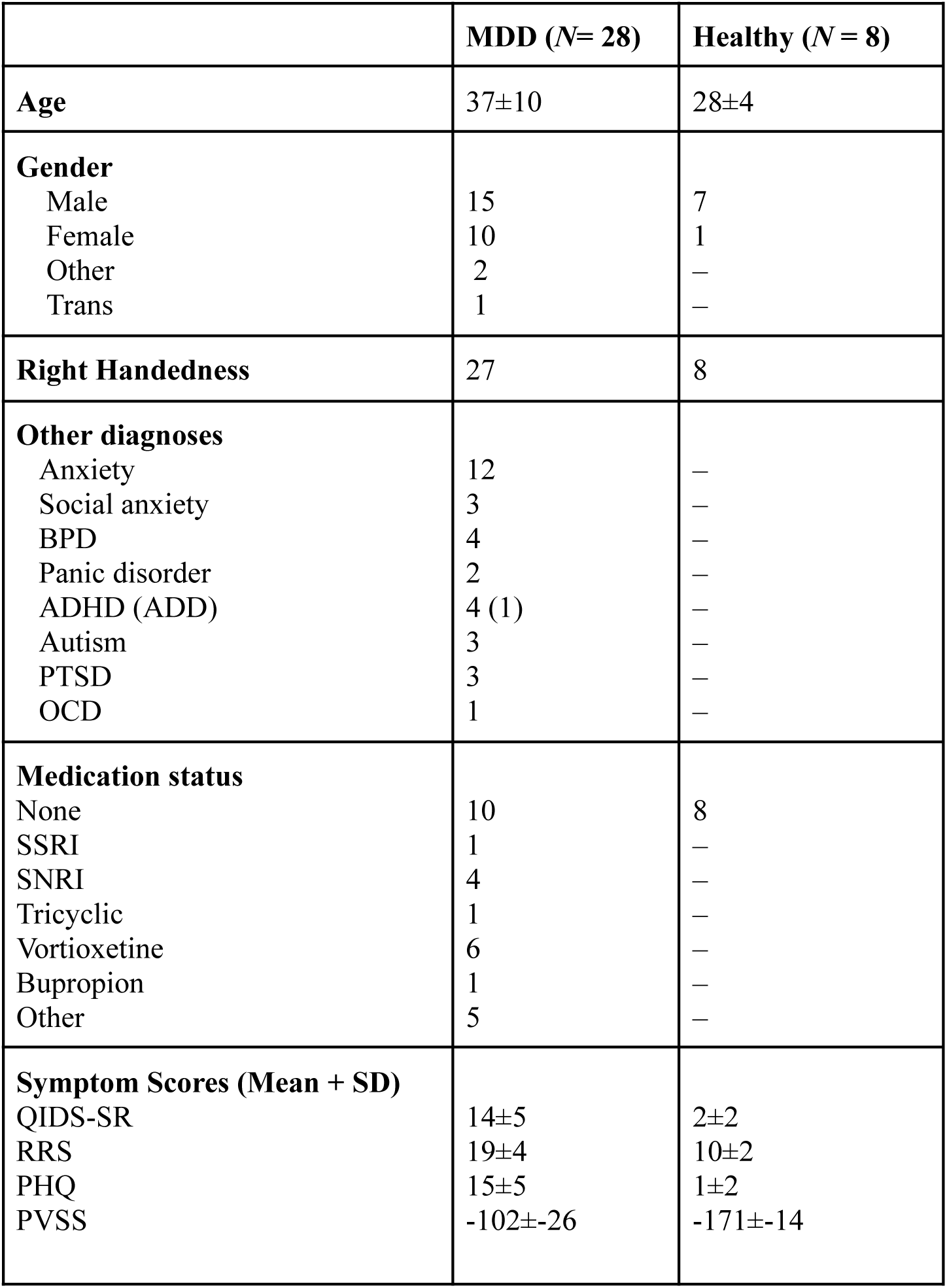
Data shown for MDD subjects (28) and healthy (8), showing mean score and standard deviation. Abbreviations: BPD, Borderline Personality Disorder, ADHD, Attention Deficit Hyperactivity Disorder, ADD, Attention Deficit Disorder, PTSD, Post-Traumatic Stress Disorder, OCD, Obsessive–Compulsive Disorder, QIDS-SR, Quick Inventory of Depressive Symptomatology Self-Report; RRS, Ruminative Response Scale; PHQ, Patient Health Questionnaire; PVSS, Positive Valence Systems Scale. PVSS scores are inverted (×−1) to match the directionality of other symptom measures.

|  | **MDD (N= 28)** | **Healthy (N = 8)** |
| --- | --- | --- |
| **Age** | 37±10 | 28±4 |
| **Gender** |  |  |
| Male | 15 | 7 |
| Female | 10 | 1 |
| Other | 2 | — |
| Trans | 1 | — |
| **Right Handedness** | 27 | 8 |
| **Other diagnoses** |  |  |
| Anxiety | 12 | — |
| Social anxiety | 3 | — |
| BPD | 4 | — |
| Panic disorder | 2 | — |
| ADHD (ADD) | 4 (1) | — |
| Autism | 3 | — |
| PTSD | 3 | — |
| OCD | 1 | — |
| **Medication status** |  |  |
| None | 10 | 8 |
| SSRI | 1 | — |
| SNRI | 4 | — |
| Tricyclic | 1 | — |
| Vortioxetine | 6 | — |
| Bupropion | 1 | — |
| Other | 5 | — |
| **Symptom Scores (Mean + SD)** |  |  |
| QIDS-SR | 14±5 | 2±2 |
| RRS | 19±4 | 10±2 |
| PHQ | 15±5 | 1±2 |
| PVSS | -102±26 | -171±14 |

Inclusion criteria for all participants were: Age 18–65, no contraindication for MRI, not pregnant or nursing, no neurological disorders such as epilepsy or brain injury.

For MDD patients additional inclusion criteria were: current major depressive episode (assessed with MINI questionnaire by a clinical coordinator, minimum inclusion more or equal to 5 points), not in danger to one’s own life, no history of psychosis, not disabled, not a prisoner nor a forensic psychiatric patient.

For healthy subjects, exclusion criteria entailed: Any diagnosis of depression or any other psychiatric disorder (assessed with MINI questionnaire), and any use of drugs or substances that affect the central nervous system (other than alcohol or nicotine products).

### MRI Acquisition

MRI data were acquired using a MAGNETOM Skyra 3T scanner (Siemens Healthcare GmbH, Erlangen, Germany) equipped with a 32-channel head coil. The MRI session included T1-weighted structural imaging, resting-state fMRI, and diffusion-weighted imaging (DWI).

### T1-Weighted Structural Imaging

T1-weighted anatomical images were obtained using a magnetization-prepared rapid gradient echo (MPRAGE) sequence with the following parameters: repetition time (TR) = 2530 ms, echo time (TE) = 3.4 ms, inversion time (TI) = 1100 ms, flip angle = 7°, field of view (FOV) = 256 x 256 mm, matrix size = 320 × 320, voxel size = 0.8 × 0.8 × 0.8 mm, and number of slices = 224.

T1-weighted images were processed with *FreeSurfer* to obtain cortical surfaces, tissue segmentations, and subject-specific parcellations. The HCP-MMP1.0 atlas (Glasser et al., 2016) was projected to individual anatomy to define cortical and subcortical ROIs.

### Resting-State Functional MRI

Resting-state functional MRI (rs-fMRI) data were collected using a T2*-weighted gradient-echo echo-planar imaging (EPI) sequence with the following parameters: TR = 1250 ms, TE = 30 ms, flip angle = 65° degrees, FOV = 126 × 126 mm, matrix size = 72 × 72, voxel size = 3 × 3 × 3 mm, number of slices = 51, and total acquisition time = 15 minutes.

Resting-state fMRI data were preprocessed with *fMRIPrep* (Esteban et al., 2019; motion and distortion correction, T1 coregistration), followed by the removal of initial volumes, denoising with motion/physiological confounds, 0.01–0.08 Hz temporal filtering, and 6-mm smoothing. Seed-to-voxel connectivity was computed using Pearson correlations (Fisher z-transformed).

### Diffusion-Weighted Imaging

Diffusion-weighted imaging (DWI) was acquired using a single-shot spin-echo echo-planar imaging (EPI) sequence. Data were collected with the following parameters: TR = 4100 ms, TE = 105 ms, FOV = 240 × 240 mm, matrix size = 80 × 80, voxel size = 3 × 3 × 3 mm, number of slices = 52, b-values = 1500, 3000 s/mm², and 64 diffusion directions. Additionally, 40 non-diffusion-weighted (b0 = 0 s/mm²) images were acquired of which 20 in reverse phase-encoding direction.

Diffusion data were preprocessed using the PyDesigner pipeline (Dhiman et al., 2024), including noise, motion, eddy-current, Gibbs, and distortion correction. Fiber orientation distributions were estimated with multi-tissue CSD and aligned to T1 space using *MRtrix3* (Tournier et al., 2019). Anatomically constrained tractography priors were derived from FreeSurfer segmentations. Real-time tractography performed with *rttvis* (Aydogan et al., 2023) was used to identify stimulation-relevant regions, prioritizing either widespread structural connectivity or dense streamline projections to network-relevant targets.

### EEG

EEG was measured with a 62-channel cap (EasyCap, Germany) with Ag/AgCl sintered electrodes connected to a TMS-compatible EEG amplifier (Brain Products, Gmbh, Germany). Signals were low-pass filtered with a 1000-Hz cut-off frequency and sampled at 5000 Hz. The reference electrode was placed on the right mastoid, and the ground over the right zygomatic bone. The skin under each electrode was gently scrubbed with abrasive conductive paste (OneStep AbrasivPlus, H + H Medical Devices, Germany) before being filled with conductive gel (Electro-Gel, ECI, Netherlands).

### TMS

TMS was applied with a 70-mm radius figure-of-eight coil attached to a TMS stimulator (NBT 2.2.4, Nexstim Ltd., Finland) with an integrated MRI-guided neuronavigation and EMG device.

### Resting motor threshold

As described in detail in Ukharova et al., 2025, to identify the RMT, single-use electrodes were placed over the right abductor pollicis brevis (APB) muscle following the belly–tendon montage. The TMS coil position and rotation was optimised to identify the target that consistently yielded the largest motor evoked potentials (MEP) from the flexion of the right APB. RMT was determined automatically by the Nexstim system’s built-in maximum-likelihood algorithm (Awiszus, 2003).

### Minimizing auditory responses to TMS

To minimize auditory confounds, noise masking was applied by playing white noise mixed with recorded TMS coil click sounds with specific time-varying frequencies of the TMS click (TAAC; Russo et al., 2022) through hybrid earplugs and earphones (ER3C Insert Earphones, Etymotic Research Inc., IL, USA).

### Target and stimulation parameters selection

For each subject, two TMS stimulation targets were selected: a standardised L-DLPFC target using Beam F3 (Beam et al., 2009), and an individualised target identified through TMS–EEG mapping using individual brain connectivity and responses to TMS stimulation.

### Standardised target

Beam F3 approximates the location of the DLPFC based on head circumference, nasion–inion distance, and left tragus to right tragus distance (Beam et al., 2009). The stimulation target was set based on these three measurements with a coil orientation of 45 degrees to the midline and stimulation intensity of 120% of the RMT (Beam et al., 2009), except 2 subjects, for whom the stimulation was delivered at 105% RMT due to discomfort and pain tolerance.

### Individualised Target

The full procedure of individualized target identification mapping is described in detail in our recent study (Ukharova et al., 2025) and summarized in Fig. 1. In brief, T1 MRIs and resting state fMRI were used to create individualized region of interest (ROI) maps, based on the functional anticorrelation between L-DLPFC and sgACC (Cash et al., 2021; Cash and Zalesky, 2024; Cole et al., 2020; Fox et al., 2012). The resulting maps were then overlaid on the individual MRI in the neuronavigation software. This ROI served as a guide for the TMS–EEG target mapping procedure prior to selecting the stimulation target within the L-DLPFC (Fig. 1A). The candidate targets were additionally investigated using real-time tractography (Aydogan et al., 2023), priority given to the cortical sites with widespread or MDD network-specific structural connectivity (Siddiqi et al., 2021).

**Fig. 1.**
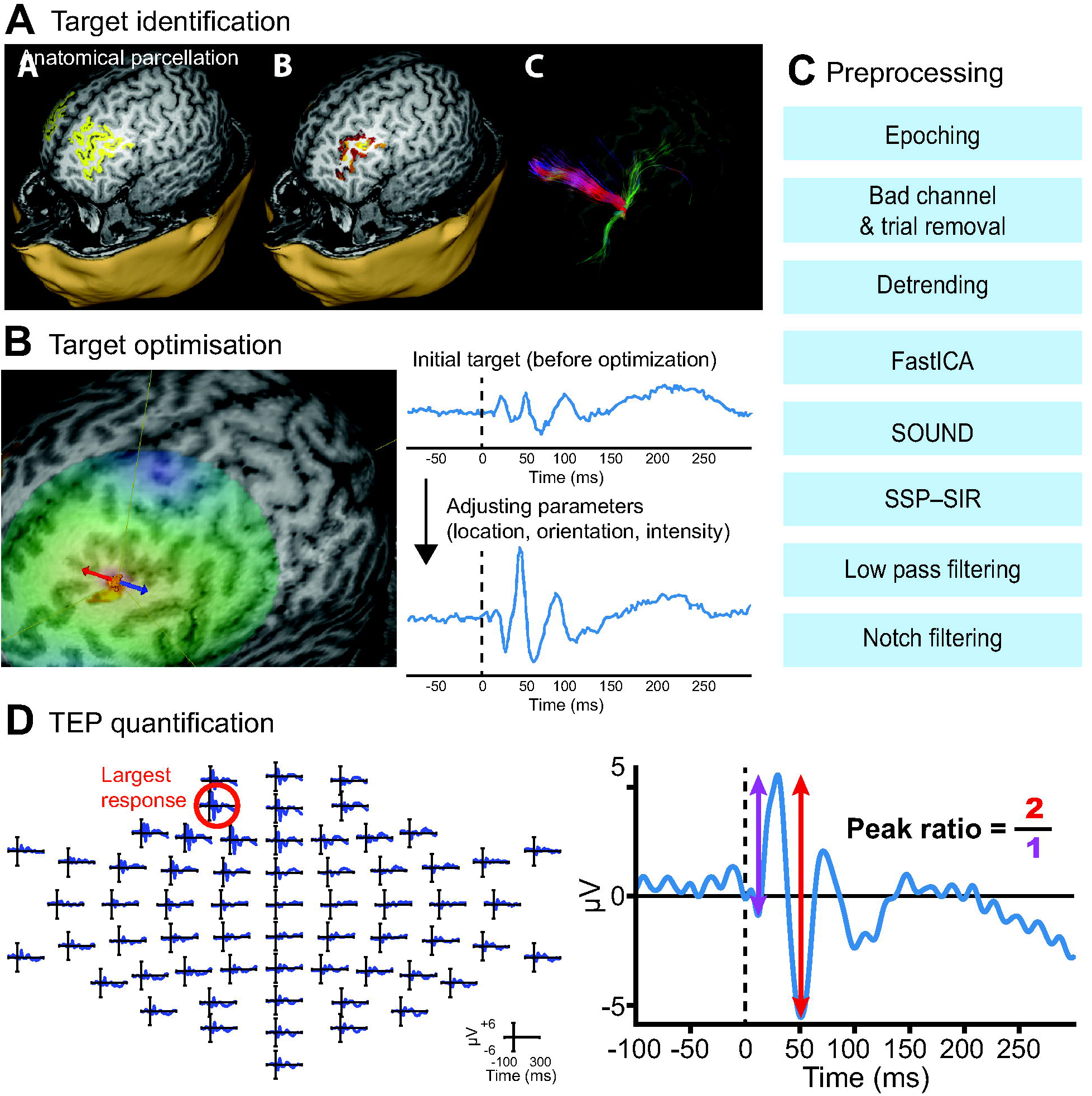
Study Schematic: A) Anatomic parcellations outlining the DLPFC region, resting-state fMRI highlighting the region of the DLPFC anticorrelated with the subgenual cingulate, lighter colours indicating greater anticorrelation, and diffusion MRI displaying the connectivity from the targeted area, all used to identify the initial individual stimulation sites for each subject. B) Target optimization using real-time E-field and TEP visualization tools (data were baseline corrected and average referenced, TMS artifact zero-padded between −2 and 10 ms). The stimulation intensity, location, and orientation were adjusted for each subject to minimise muscle artefacts and obtain a 6–10 µV early response (10–60 ms). C) The data preprocessing pipeline, described in more detail by Mutanen et al. (2024). D) TEP quantification from an electrode with the most pronounced response for each subject (one subject shown as an example). The ratio of the second peak-to-peak over the first peak-to-peak was calculated for each subject, and correlated with depressive symptom scores (PHQ-9, RRS, PVSS, QIDS-SR).

The coil was initially placed over one of the areas displaying the highest functional anticorrelation with sgACC within the L-DLPFC, in the posterior-anterior direction of the induced electric field, and stimulus intensity was set to 100–110% of RMT over this initial location. Adjustments were made based on two criteria: a) elimination of artifacts after the first 10 ms, b) a 6–10 μV peak-to-peak response of early TEP deflections (10–60 ms). If a muscle artifact was present with a duration exceeding 10 ms, the coil was rotated and/or coil position manipulated towards an artifact-free location, while staying within the L-DLPFC defined by the individual anatomical parcellation (for details, see Ukharova et al., 2025). Stimulation intensity was adjusted in 2% increments of the maximum stimulator output until the desired peak-to-peak amplitude was obtained (Supplementary Fig. 1). The final stimulation site was chosen based on both its connectivity profile and the quality of the TMS–EEG response (Lioumis and Rosanova, 2022).

### Recordings

Prior to and during the recording, electrode impedances were kept below 5 kΩ. At each target, 300 single pulses were delivered with a jittered interstimulus interval of 2–2.3 s.

### EEG data preprocessing

The recorded EEG data were preprocessed in MATLAB R2017b using the EEGLAB toolbox (Delorme and Makeig, 2004) with in-house custom scripts. The recorded EEG signals were segmented into epochs of −1500 to 1500 ms around the TMS pulse. Baseline correction was applied with a baseline of −500 to −5 ms. To remove the TMS pulse artifact, data from −2 to 7 ms were removed and interpolated with a cubic spline. Noisy channels and trials with heavily distorted components were removed manually. Drifts were suppressed with robust detrending. Ocular artefacts and continuous muscle activity were removed with independent component analysis (FastICA; Oja and Yuan, 2006). Noise from extracranial sources was suppressed with the source-estimate-utilizing noise-discarding algorithm (SOUND; Mutanen et al., 2018); any remaining TMS-evoked muscle artefacts were suppressed with signal-space projection–source-informed reconstruction (SSP–SIR; Mutanen et al., 2016). Low-pass filtering was applied with a cut-off frequency of 80 Hz and notch filtering at 48–52 Hz. For statistical analysis, the epochs were reduced to −100 to 300 ms.

### Quantification of TMS-evoked potentials (TEPs)

For each subject, TMS-evoked potentials (TEPs) were averaged across trials for each channel of interest within the L-DLPFC (AF3, F1, F3, F5, FC1, FC3). Peaks were identified as local maxima and minima using a minimum prominence criterion of 0.2 μV (Fig. 1D). The search window spanned 10–60 ms post-stimulation (extended to 70 ms in one subject in which fewer than three peaks were detected in the 10-60 ms window). For each channel, the first three peaks in time were extracted, and the maximum absolute amplitude difference between consecutive peaks was computed. The channels were ranked based on the largest consecutive peak amplitude difference among these three peaks. Subsequently, all selected channels were visually inspected to ensure that the channels exhibiting the most pronounced TEP morphology were retained (Supplementary Table 1). When two peaks appeared within 10–20 ms, the later peak was treated as the first to minimize residual artefacts and capture genuine early TEP activity.

Subsequently, peak ratio was calculated as

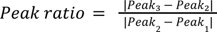, where each 𝑃𝑒𝑎𝑘 represents the amplitude of the corresponding peak.

Additionally, N100 peak was identified as the local maximum within the 80–140-ms window. Peak latencies, amplitudes, and inter-peak differences were recorded for further analyses (Table 2, Supplementary Figure 2).

**Table 2.** Mean amplitudes (μV±SD) and latencies (ms±SD) of TMS evoked potentials in healthy (*N* = 8) and MDD subjects (*N* = 28) in the ipsilateral (left) hemisphere at the individualised L-DLPFC target.

|  |  | Peak 1 | Peak 2 | Peak 3 | N100 |
| --- | --- | --- | --- | --- | --- |
| Healthy | Amplitude<br>Latency | 0.93±2.81<br>14.19±3.48 | -1.61±5.12<br>25.26±6.54 | 1.95±1.52<br>41.46±8.32 | -2.25±1.11<br>91.78±13.14 |
| MDD | Amplitude<br>Latency | -1.52±6.42<br>15.48±3.95 | 0.19±3.34<br>27.13±9.43 | 0.58±0.21<br>39.93±14.13 | -1.3±1.03<br>102.42±19.4 |

### Questionnaires

Primary symptom scores were measured with the Patient Health Questionnaire (PHQ-9). Secondary symptom scores were evaluated using the Quick Inventory of Depressive Symptomatology Self-Report (QIDS-SR), Ruminative Response Scale (RRS), and Positive Valence Symptom Scale (PVSS). PVSS scores were inverted (×−1) to match the directionality of other symptom measures.

### Statistical analysis

All statistical analyses were performed in Python (v3.12.11) using *pandas*, *numpy*, *scipy*, and *statsmodels* libraries. Data distribution was assessed with histograms and Shapiro–Wilk tests. As the data were not normally distributed, non-parametric statistical analyses were conducted. Mann–Whitney U-test was performed to compare the peak ratios between groups. Spearman rank correlations were used to assess relationships between the peak ratio (second peak-to-peak amplitude over the first peak-to-peak amplitude) and depression symptom scores. The *p*-values for correlations with questionnaire scores were corrected for multiple comparisons using the Benjamini–Hochberg false discovery rate (FDR) method; FDR-corrected values are reported as *q*-values. To evaluate whether antidepressant medication influenced the group differences in peak ratios, we conducted a sensitivity analysis restricted to unmedicated participants. Group differences were assessed using Welch’s *t*-test, given the reduced sample, with effect sizes quantified using Hedges’ *g*. The results were additionally confirmed using the non-parametric Mann–Whitney U test.

## Results

### Demographics and Clinical Characteristics

The study sample consisted of 28 MDD patients and 8 healthy controls. A summary of the demographics of study participants is available in Table 1. The MDD group included a range of comorbid conditions and medication status.

### Peak ratio and N100 amplitude reflect behavioral scores in MDD

To study how the TMS–EEG evoked potentials are related to the MDD severity, we correlated the N100 amplitude with the total scores of the PHQ, QIDS-SR, RRS, and PVSS scores. The Mann–Whitney U test revealed a significant difference in peak ratio for the mapped target between healthy (*N* = 8) and MDD (*N* = 28) participants (*U* = 34, *p* = 0.002, *Z* = −2.968, *r* = 0.495), indicating a moderate effect. No group difference was found between groups for peak ratios derived from TEPs recorded over the standardized Beam F3 target.

To assess potential medication effects, we restricted the sample to unmedicated participants with MDD. The group difference in peak ratio remained significant (healthy: *N* = 8; unmedicated MDD: *N* = 10; Welch’s *t*-test (*t*=−3.26, *p* = 0.009; Hedges’ *g* = 1.38), confirmed using a non-parametric Mann–Whitney U test (*U*=0, *p* < 0.001), indicating robustness to medication status.

For the individualised mapped L-DLPFC targets (Fig. 2), peak ratio correlated with QIDS-SR (*ρ* = 0.459, *q* = 0.01), RRS (*ρ* = 0.656, *q* = 6×10⁻⁵), PHQ (*ρ* = 0.368, *q* = 0.037), and PVSS (*ρ* = 0.332, *q* = 0.048). Higher ratios were associated with greater symptom severity.

**Fig. 2.**
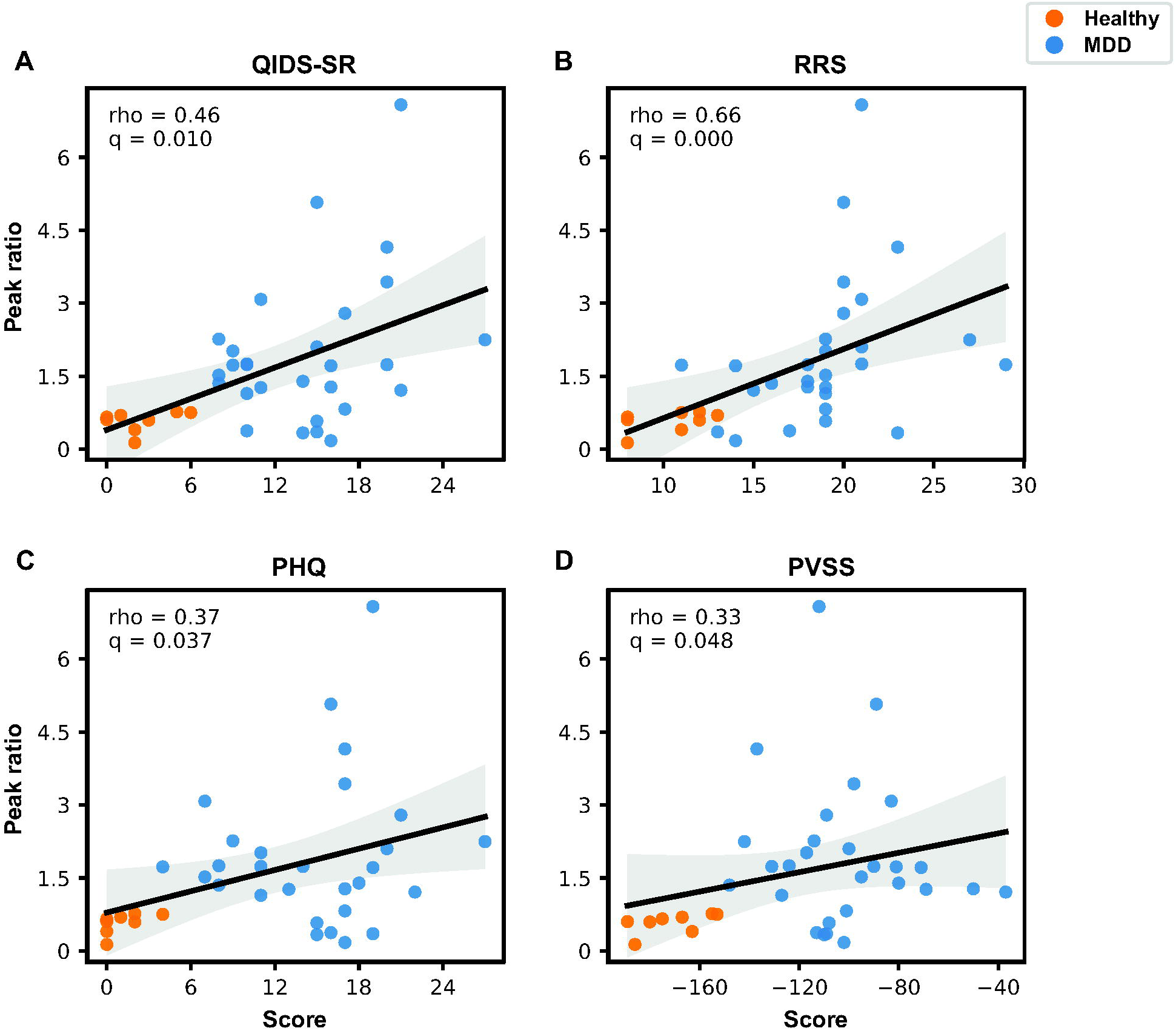
Scatter plots illustrating the association between peak ratio measured at the individualized mapped L-DLPFC target and symptom severity scores (QIDS-SR [A], RRS [B], PHQ [C], PVSS [D]) in healthy and MDD participants. Black lines show the pooled linear regression fit with shaded 95% confidence intervals. Spearman correlation coefficients and corresponding *q*-values reflect analyses across all participants combined.

For the Beam F3 targets (Fig. 3), peak ratio was not significantly correlated with any symptom scores: (QIDS-SR: *ρ* = 0.018, *q* = 0.919; RRS: *ρ* = 0.065, *q* = 0.919; PHQ: *ρ* = 0.050, *q* = 0.919; PVSS: *ρ* = 0.102, *q* = 0.919).

**Fig. 3.**
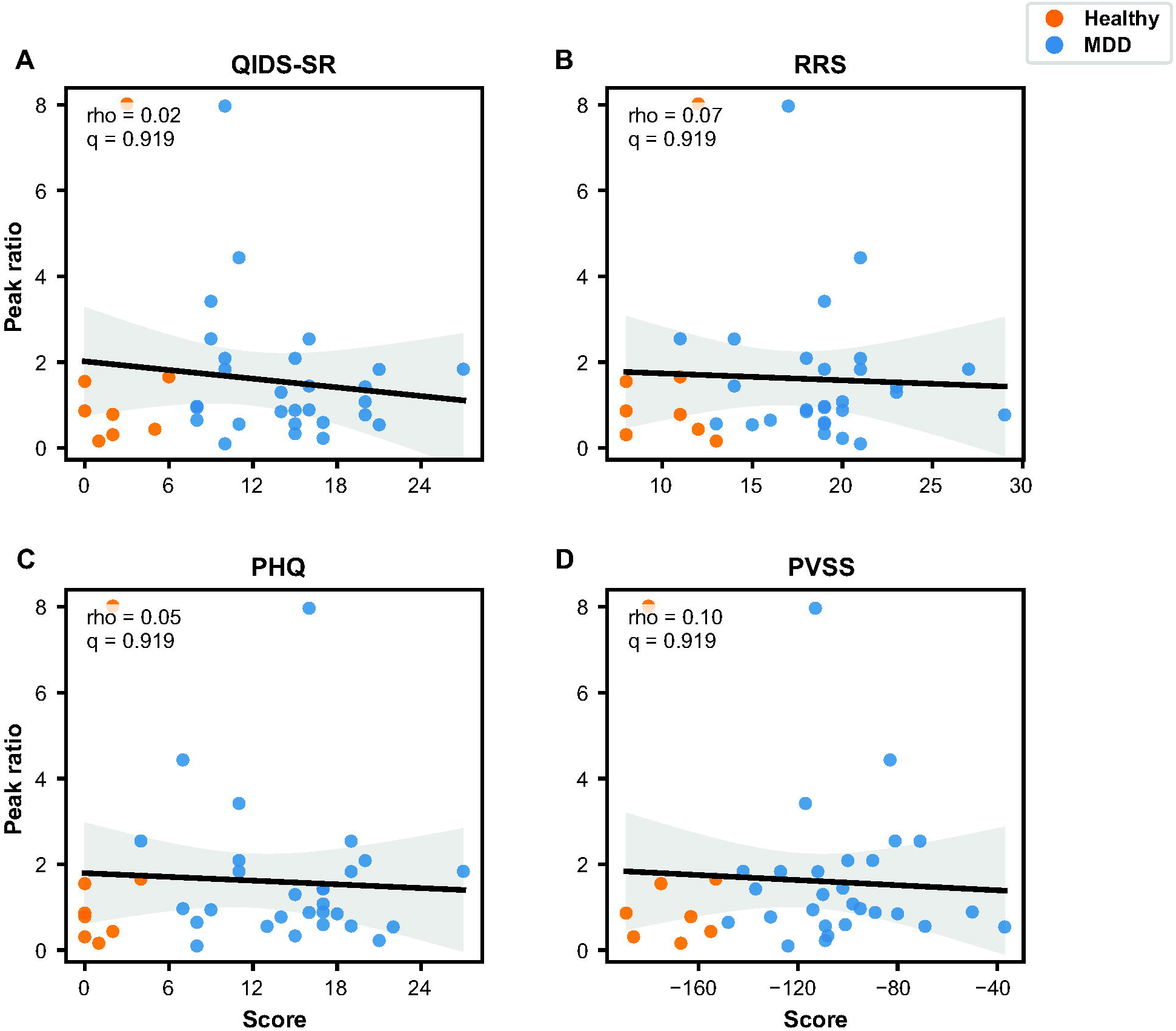
Scatter plots illustrating the association between peak ratio measured at the Beam F3 target and symptom severity scores (QIDS-SR [A], RRS [B], PHQ [C], PVSS [D]) in healthy and MDD participants. Regression lines (black) and 95% confidence intervals are shown. Spearman correlation coefficients and *q*-values correspond to pooled analyses including all participants.

At the mapped target (Fig. 4), N100 amplitude correlated with QIDS-SR (*ρ* = 0.445, *q* = 0.012), RRS (*ρ* = 0.355, *q* = 0.034), PHQ (*ρ* = 0.391, *q* = 0.025), and PVSS (*ρ* = 0.469, *q* = 0.013), whereas N100 latency showed no associations. At Beam F3 target, neither N100 amplitude nor latency correlated with symptom scores.

**Fig 4.**
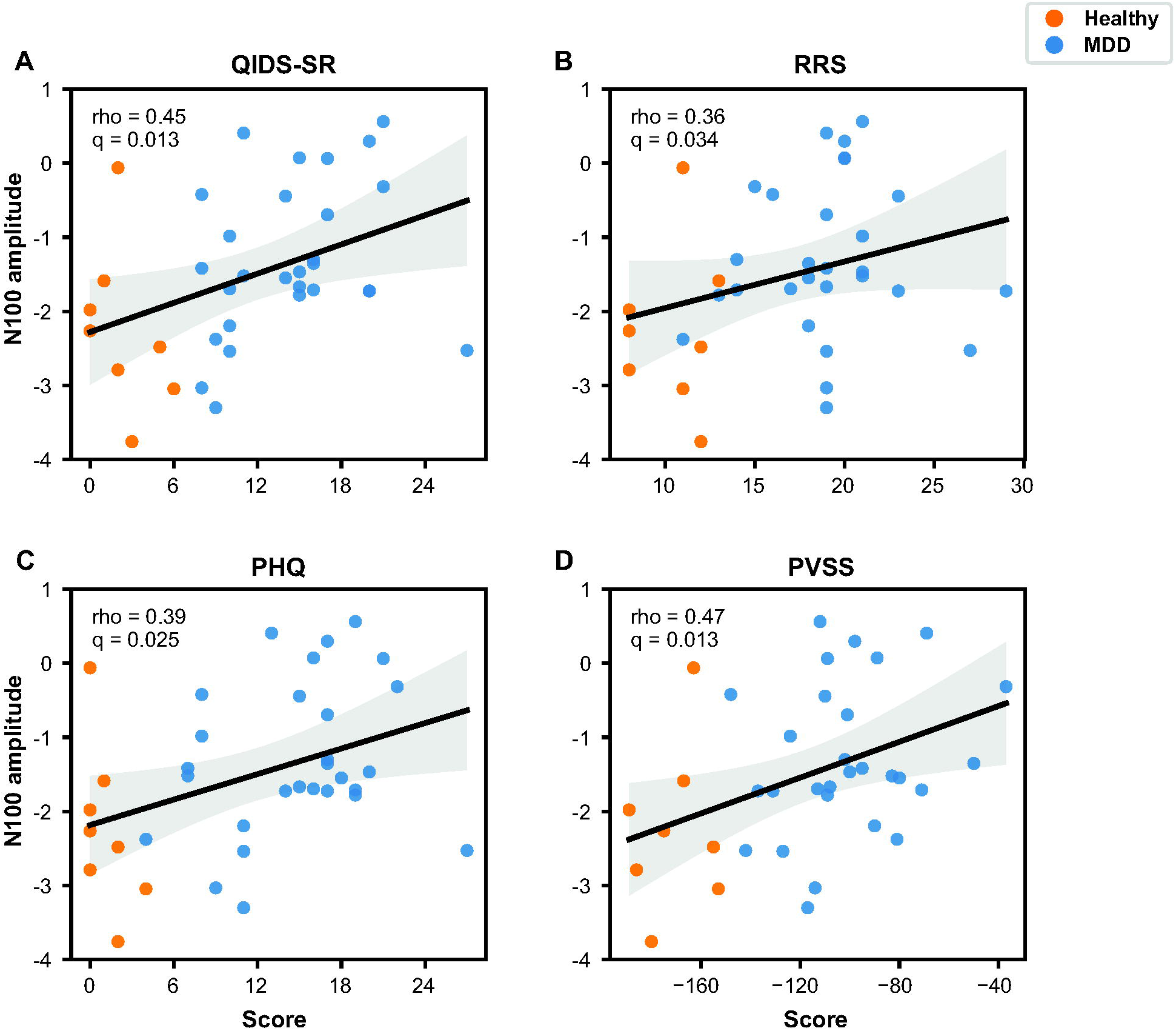
Scatter plots depicting the relationship between N100 amplitude derived from the individualized mapped L-DLPFC target and symptom severity scores (QIDS-SR [A], RRS [B], PHQ [C], PVSS [D])for healthy and MDD participants. Black regression lines indicate the pooled fit with 95% confidence intervals. Reported Spearman correlation coefficients and *q*-values are computed across all participants.

### TMS-evoked potentials in MDD and healthy populations

TMS to the L-DLPFC elicited canonical TEP components (N15, P30, N45, P60, N100) at the group level (Table 2), although peak expression varied across individuals (Supplementary Fig. S2). As shown in Fig. 5, early responses were larger at individualised targets than Beam F3. Peak latencies did not differ between MDD and healthy participants (Table 2).

**Fig. 5.**
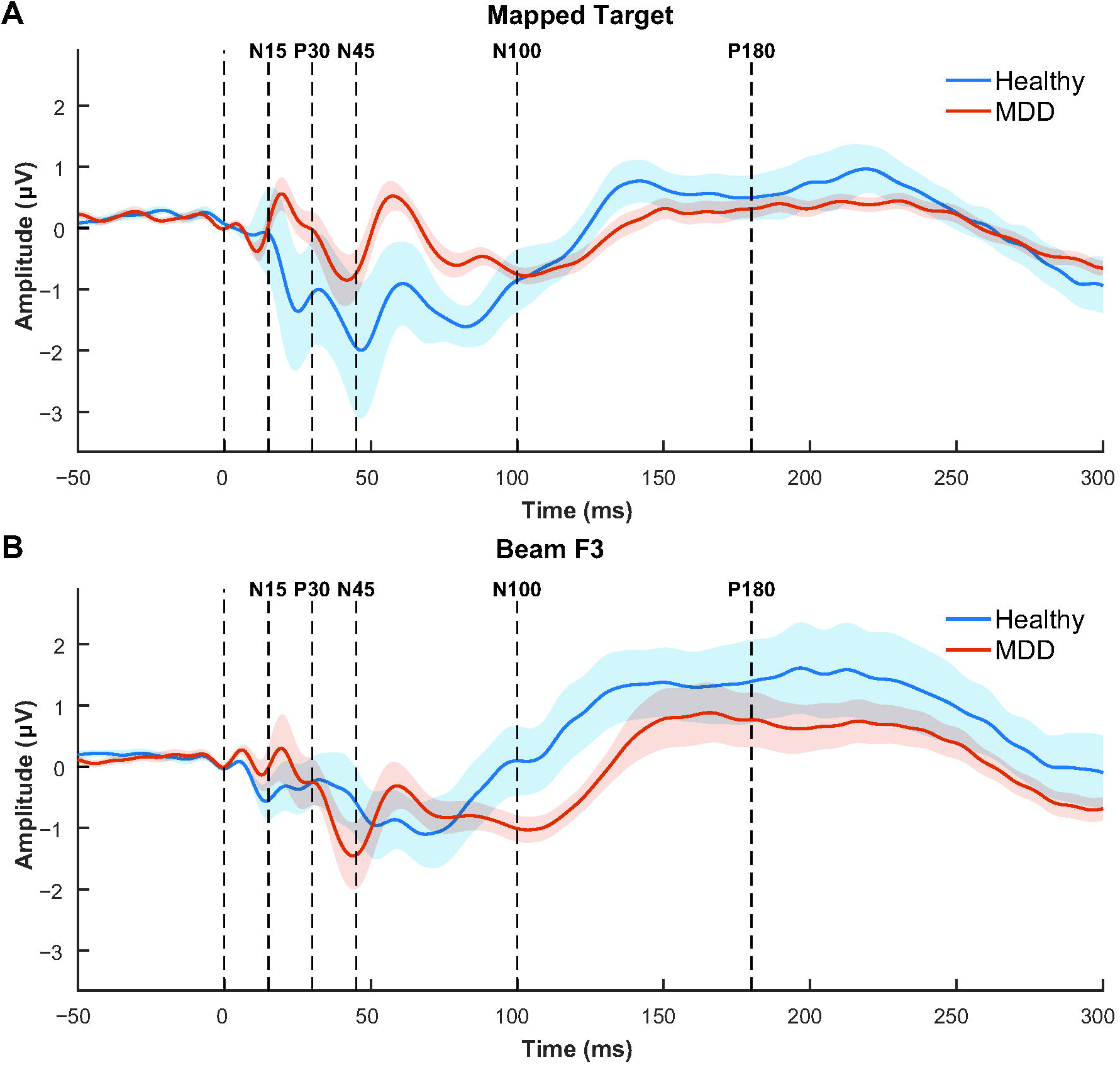
Grand-average TEP waveforms from the prefrontal ROI (AF3, F1, F3, F5, FC1, FC3) from −100 to 300 ms following the TMS pulse in healthy participants (*N* = 8) and individuals with MDD (*N* = 28). A) Stimulation over individualized mapped target. B) Stimulation at Beam F3.

Group-average butterfly plots and scalp topographies at individualized targets are shown in Fig. 6.

**Fig. 6.**
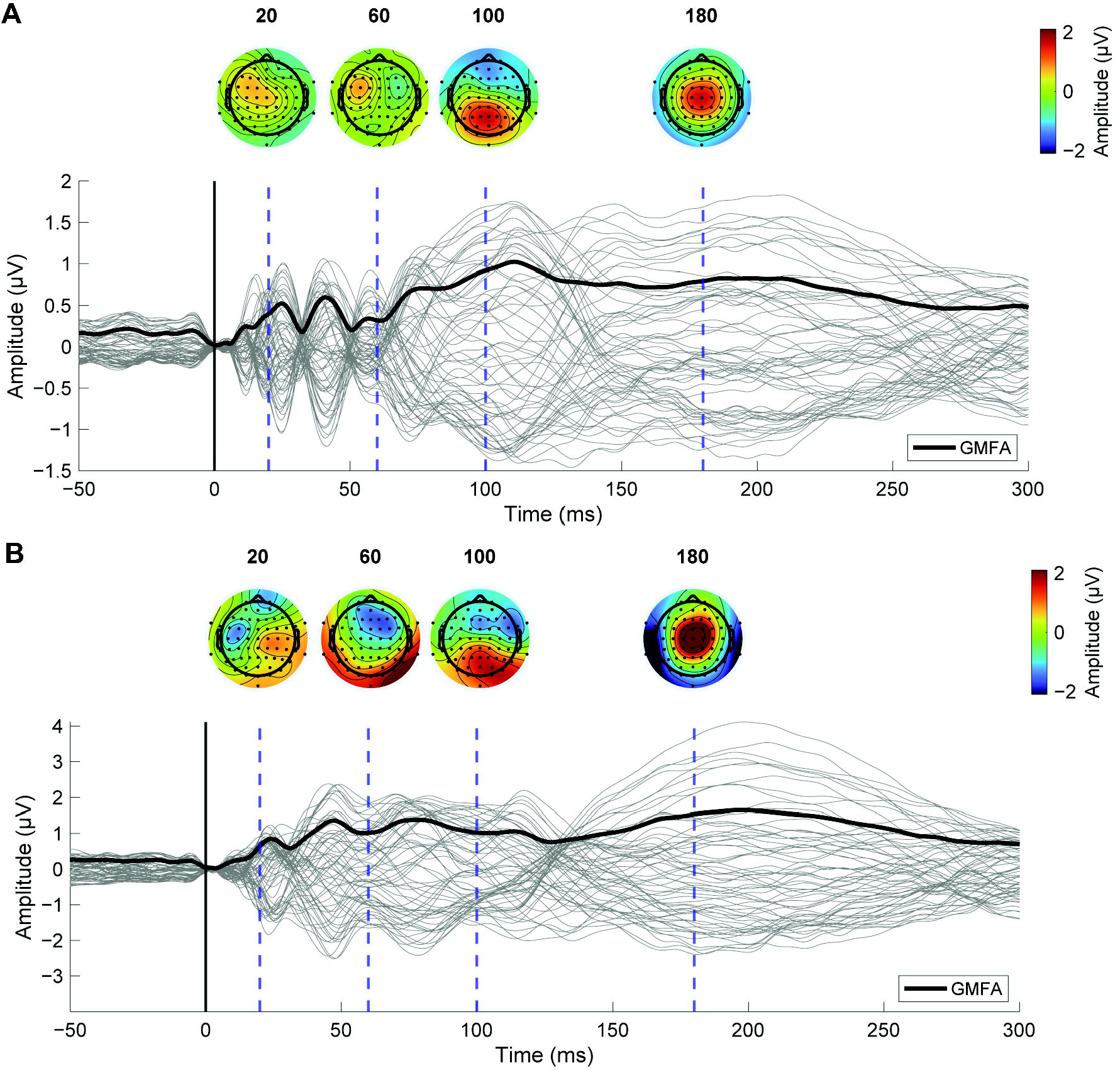
Butterfly plots and scalp topographies for 38 MDD participants (A) and 8 healthy participants (B) during stimulation of the mapped L-DLPFC target. The black vertical line marks the TMS pulse onset (0 ms). Blue dashed vertical lines at 20, 60, 100 and 180 ms indicate the time points corresponding to the displayed scalp topographies.

## Discussion

This study demonstrates that individualized targeting of the L-DLPFC enables reliable measurement of early TMS–EEG responses that scale with depressive symptom severity, an effect not observed with Beam F3 targeting. These findings suggest that tailoring stimulation to individual head and brain morphology is critical for minimizing artifacts and accessing early TEP components as sensitive indices of depression-related neurophysiology, particularly in small, heterogeneous samples.

We compared sensitivity of TEPs recorded using individualized mapping versus the standard Beam F3 stimulation protocol at 120% of the RMT. Early TEP responses elicited at Beam F3 were not predictive of clinical symptom severity. This may reflect either suboptimal stimulation location or stimulation intensity. In the first case, low local cortical excitability may have produced insufficient neural responses, or the targeted region may have been heavily contaminated by muscle artifacts (Gogulski et al., 2024b), the removal of which inevitably attenuates genuine cortical activity. In the second case, inadequate stimulation intensity—either too low to effectively perturb the cortex or too high, thereby amplifying muscle artifacts—could similarly reduce the sensitivity of TEP measures. These results highlight the necessity of individualized cortical mapping for developing reliable TMS–EEG-based biomarkers of MDD.

Prior biomarker research has focused on later TEP components, such as the N100 and P200, owing to their high reproducibility due to the lack of stimulation-related artifacts (Cao et al., 2021; Kallioniemi and Daskalakis, 2022; Sun et al., 2018, 2016). However, later responses are affected by auditory and somatosensory responses to the stimulation, as well as signal propagation confounds (Conde et al., 2019; Ross et al., 2022), limiting their specificity for local cortical reactivity (Hernandez-Pavon et al., 2023; Tremblay et al., 2019). In contrast, early components (<60 ms) may more directly reflect local cortical responses and dynamics (Casarotto et al., 2022; Ilmoniemi et al., 1997; Tremblay et al., 2019), but are prone to being masked by muscle and peripheral nerve artifacts, leading to lower reliability unless the signal-to-noise ratio is actively optimized (Lioumis and Rosanova, 2022). Recent methodological advances emphasize real-time artifact minimization and EEG-guided targeting as key strategies for recovering early responses (Casarotto et al., 2022; Tervo et al., 2022; Varone et al., 2025).

Consistent with these recently introduced approaches, our individualized EEG-guided mapping allowed to record artifact-reduced early TEPs in the DLPFC that revealed symptom correlations absent with Beam F3 targeting, suggesting the increased sensitivity. These findings align with evidence that optimized DLPFC targeting increases early responses and reduces muscle contamination (Gogulski et al., 2024a; Parmigiani et al., 2025; Ukharova et al., 2025). These results support the clinical feasibility of using individualized early TEP components as informative readouts of prefrontal neurophysiological activity.

We quantified local cortical activity using a ratio of peak-to-peak amplitudes between 10–60 ms, normalizing across individualized targets and intensities. As specific TEP components have been linked to both excitatory and inhibitory neurotransmitters, we interpret this ratio as an index of local excitation–inhibition (E/I) balance. The earliest peaks, N15 and P30, have been suggested to be modulated by NMDA antagonists, while the N45 and P60 components have been linked to GABA-A-ergic activity (Belardinelli et al., 2021; Premoli et al., 2014; Salavati et al., 2018). As such, a larger second peak-to-peak observed in MDD patients may reflect increased inhibition, whereas the opposite pattern may indicate decreased inhibitory feedback. However, further pharmacological and intracranial validation is essential for refining the interpretations of TEP-based biomarkers.

Our results are consistent with previous literature on TMS–EEG biomarkers in MDD, which reported abnormalities in inhibitory components (N45, N100) across multiple cohorts, supporting evidence of impaired GABAergic transmission in depression (Darmani and Ziemann, 2019). We also demonstrated statistically significant correlation of the behavioural scores with the amplitude of the N100 for the mapped target, confirming the importance of N100 as an MDD biomarker. However, we did not observe a similar effect with Beam F3 targeting as in previous studies, likely due to the small size of the study population.

It is important to note that individualized EEG-based mapping yields targets optimized for monitoring and biomarker purposes rather than treatments; locations allowing for reliable recording of the early TEPs will not necessarily produce maximal clinical benefit. Nonetheless, the amplitude ratio of the early TEPs offers an objective, physiologically interpretable biomarker and a practical method to define the stimulation intensity for rTMS therapy beyond RMT heuristics (see Supplementary Fig. 1). Current dosing approaches generally utilise the RMT, which reflects M1 excitability and may not adequately predict the responsiveness of the prefrontal cortex (Chang et al., 2025; Tik et al., 2025). Cortical, site-specific early TEPs could help titrate the intensity required to perturb DLPFC in an individual patient, warranting prospective studies directly comparing RMT- versus TEP-guided dosing.

Several limitations should be noted. Individualizing stimulation parameters improves within-subject interpretability but complicates between-subject comparisons. Longitudinal studies are necessary to determine whether the peak ratio reflects symptom change over time. In addition, the clinical heterogeneity of the patient group (in terms of medication and comorbidities) may have influenced the observed effects. More homogeneous and medication-naïve cohorts could yield clearer insights into the neurophysiological mechanisms underlying peak-amplitude differences. Finally, while the observed E/I imbalance may be associated with depressive symptomatology, additional studies are required to clarify whether it represents a causal mechanism.

A critical consideration for the clinical translation of our method is that manual mapping of the DLPFC is time-consuming and highly dependent on the operator’s expertise and the available hardware and real-time monitoring setup; it is often complicated by the presence of artifacts (Lioumis and Rosanova, 2022). Translating individualized mapping for targeting and stimulation intensity selection into clinical practice will require automation of the mapping and stimulation procedures to ensure reproducibility and operator-independent implementation, consistent with emerging automated frameworks described by Parmigiani et al. (2025) and Tervo et al. (2022) that can be applied with the emerging technology (Matsuda et al., 2025; Nieminen et al., 2022; Sinisalo et al., 2024). Importantly, despite the current limitations, our experimental setup demonstrates for the first time the feasibility of extracting real-time biomarkers during DLPFC stimulation.

Individualized DLPFC mapping enables reliable measurement of artifact-reduced early TEPs, providing a quantification method that captures early excitatory and inhibitory dynamics through a ratio of peak-to-peak amplitudes that scales with depressive symptom severity. Our approach may not only provide a physiologically grounded biomarker for MDD, but also introduce a method to calibrate prefrontal stimulation intensity beyond RMT heuristics, potentially increasing response rates due to sufficient stimulation.

## Data Availability

Data produced in the present study are available upon reasonable request to the authors.

## Acknowledgements

We would like to acknowledge the contribution of the MDD patients and healthy volunteers that led to the development of our experimental protocol.

This work was supported by Wellcome Leap as part of the Multi-Channel Psych Program. This project has also received funding from the European Research Council (ERC) under the European Union’s Horizon 2020 Research and Innovation Programme (grant agreement No 81037).

JG has received a personal grant from Emil Aaltonen Foundation and Young Group Leaders Grant from Sigrid Juselius Foundation, supporting work of JG and CT.

