## Supplementary materials for "TMS–EEG-derived excitation/inhibition ratio as a diagnostic biomarker for major depressive disorder"

Affiliations:
[1]Department of Neuroscience and Biomedical Engineering, Aalto University School of Science, Espoo, Finland
[2]Department of Clinical Neurophysiology, HUS Diagnostic Center, Helsinki University Hospital and University of Helsinki, Helsinki, Finland
[3]BioMag Laboratory, HUS Medical Imaging Center, Aalto University, University of Helsinki and Helsinki University Hospital, Helsinki, Finland
[4]Division of Psychology, VISE, Faculty of Education and Psychology, University of Oulu, Finland
[5]Neuroscience Center, Helsinki Institute of Life Science, University of Helsinki, Finland
[6]Department of Psychiatry, University of California San Diego, San Diego, California
[7]Temerty Centre for Therapeutic Brain Intervention, Centre for Addiction and Mental Health
[8]Department of Psychiatry, Temerty Faculty of Medicine, University of Toronto
[9]Department of Clinical Neurophysiology, HUS Diagnostic Center, Helsinki University Hospital and University of Helsinki, Helsinki, Finland
[10]Cognitive Brain Research Unit, Department of Psychology and Logopedics, Faculty of Medicine, University of Helsinki, Helsinki, Finland
[11]School of Psychology and Neuroscience, University of Glasgow, Glasgow, United Kingdom

†These authors contributed equally
*Corresponding author

Keywords: Major depressive disorder, depression, transcranial magnetic stimulation, TMS–EEG, cortical excitability, biomarkers

### Supplementary Results

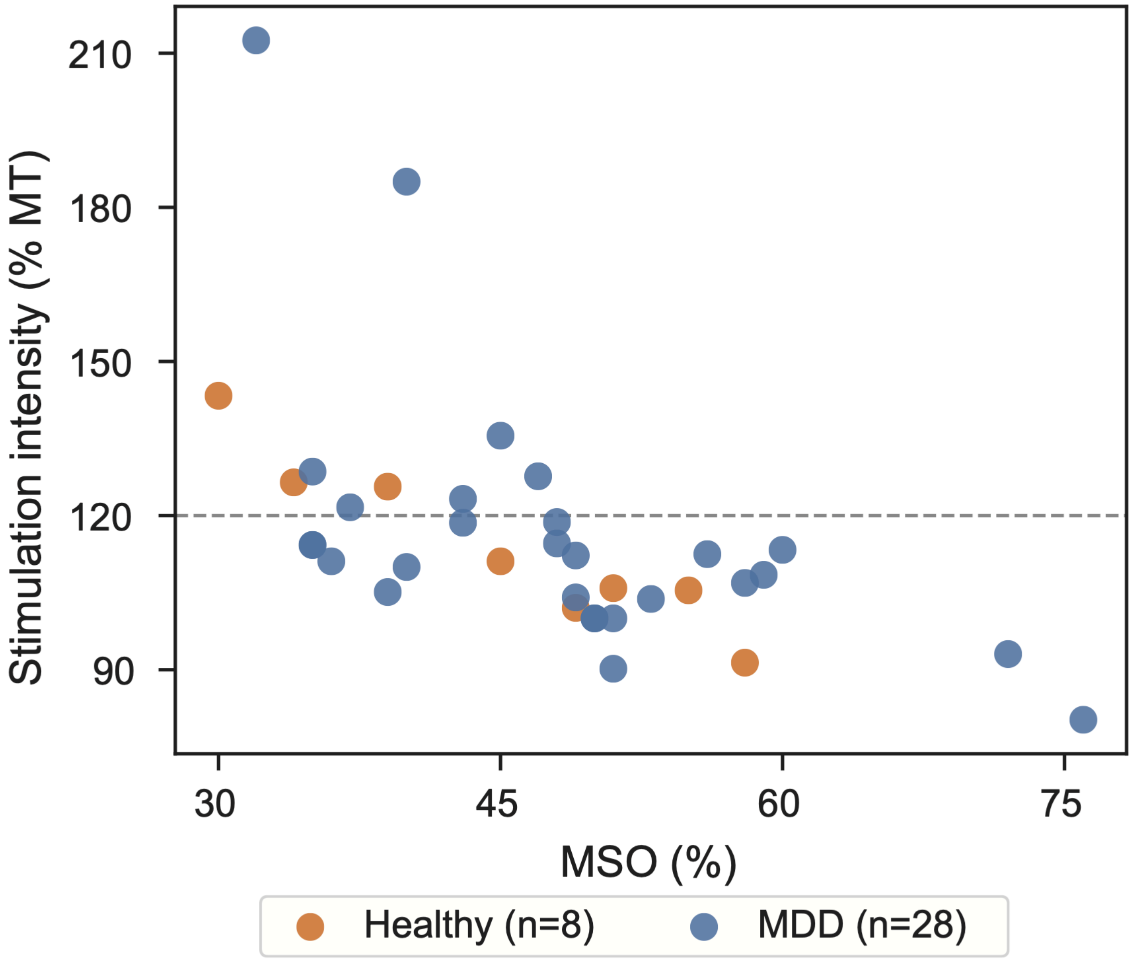


Fig. S1. Scatter plot of stimulation intensity relative to individual RMT for healthy and MDD participants at the mapped L-DLPFC target. The dashed grey line at 120% RMT denotes the commonly used stimulation level for Beam F3 targeting.

| **Electrode** | **Number of subjects** |
| --- | --- |
| AF3 | 6 (6 MDD, 2 HC) |
| F1 | 10 (5 MDD, 3 HC) |
| F3 | 6 (7 MDD, 2 HC) |
| F5 | 1 (2 MDD) |
| FC1 | 9 (4 MDD) |
| FC3 | 10 (4 MDD, 1 HC) |

Supplementary Table 1: Electrodes used for TEP peak quantification

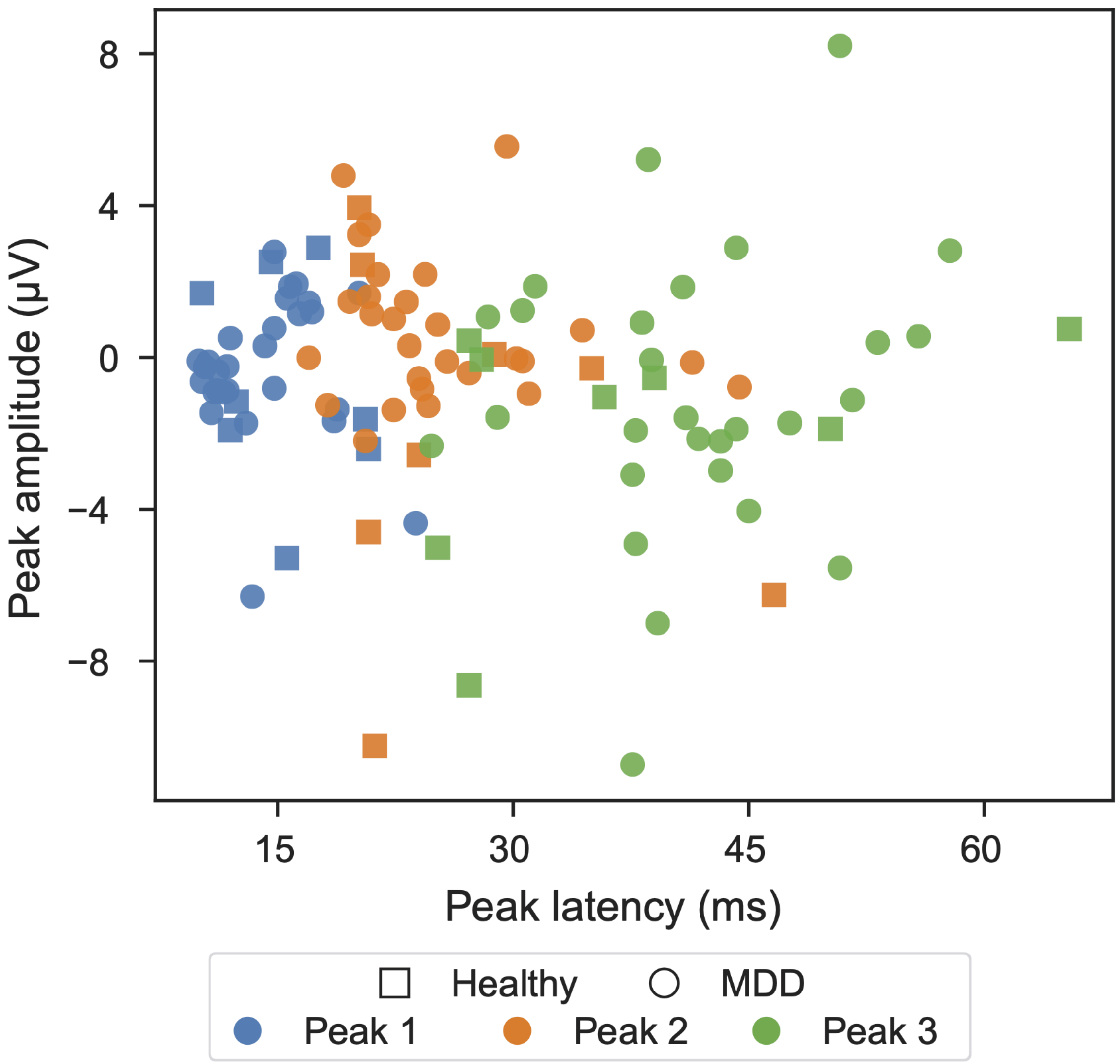


Fig. S2. Scatter plot of peak latency (ms) and peak amplitude (μV) across 28 MDD and 8 healthy participants.

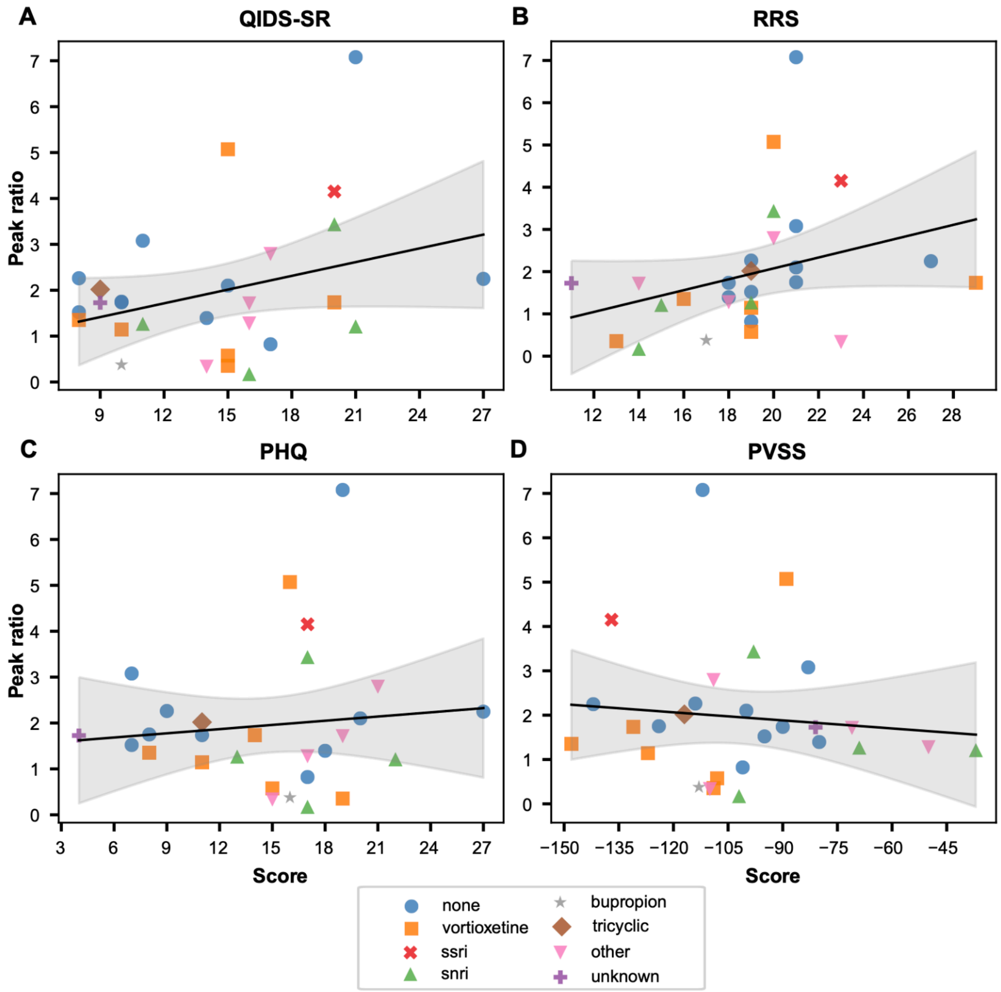


Fig. S3. Scatter plots showing the relationship between the ratio of the second to first peak amplitudes at the mapped L-DLPFC target and symptom scores (QIDS-SR [A], RRS [B], PHQ [C], PVSS [D]) for MDD participants only. Marker shape and colour denote medication status.
